# Association of Medicaid Expansion Under the Affordable Care Act with Insurance Status and Clinical Characteristics of Low-Income Patients with Newly Diagnosed Melanoma

**DOI:** 10.1101/2020.08.22.20179903

**Authors:** Pranav Puri, Mark R. Pittelkow, Lanyu Mi, Aaron R. Mangold

**Author notes:** **Correspondence:** Pranav Puri, 13400 E Shea Blvd, Scottsdale, AZ 85259 **Email:**. **Funding sources:** None. **IRB approval status:** Not applicable due to publicly available data.

## Abstract

**Importance:** The Affordable Care Act expanded Medicaid eligibility in participating states to individuals with incomes up to 138% of the federal poverty line. The effects of this policy on the diagnosis and treatment of melanoma in low-income populations has yet to be described.

**Objective:** To evaluate the effect of Medicaid expansion on changes in insurance status and clinical characteristics of low-income patients with newly diagnosed melanoma.

**Design, Setting, and Participants:** This cross-sectional study included patients younger than 65 with a new diagnosis of malignant melanoma from January 1, 2011 to December 31, 2016, in the US National Cancer Institute’s Surveillance Epidemiology and End Results database.

**Exposures:** Residence in a state that expanded Medicaid on January 1, 2014.

**Main Outcomes and Measures:** The primary outcomes were insurance status, melanoma staging, and overall survival.

**Results:** In Medicaid expansion states, there were 1,719 low-income patients with newly diagnosed melanoma during the pre-expansion time period and 1,984 (15% increase) during the post-expansion time period. In nonexpansion states, there were 326 low-income patients with newly diagnosed melanoma during the pre-expansion time period, and 288 during the post-expansion time period (12% decrease). Compared with nonexpansion states, expansion states had a significantly greater reduction in percentage of uninsured patients following Medicaid expansion (adjusted odds ratio, 6.27 [95% CI, 4.83 to 8.14]). Overall survival was not statistically different between expansion and nonexpansion states (HR, 0.89 [95% CI, 0.74 to 1.06]). There were no statistically significant differences in melanoma staging at diagnosis between the expansion and nonexpansion groups (p = 0.05).

**Conclusions and Relevance:** Medicaid expansion was associated with increased melanoma diagnoses in low-income patients and a decreased proportion of uninsured patients. However, our study did not identify differences in clinical outcomes associated with Medicaid expansion.

**Key Points:** *Question:* Was Medicaid expansion associated with changes in insurance status and clinical characteristics of low-income melanoma patients?

*Findings:* Medicaid expansion was associated with increased diagnoses of melanoma in low-income populations and reductions in the proportion of uninsured melanoma patients. However, there were no statistically significant changes in staging at diagnosis or overall survival associated with Medicaid expansion.

*Meaning:* Increased health insurance coverage associated with Medicaid expansion could potentially improve timely detection and treatment of melanoma for low-income populations.

## Introduction

The Affordable Care Act (ACA) included provisions and federal funding for states to expand Medicaid eligibility to adults earning up to 138% of the federal poverty line. Yet, in 2014, only 24 states accepted federal funding to expand their Medicaid programs. In these states, Medicaid expansion resulted in millions of low-income adults gaining insurance coverage.^1^ In addition, Medicaid expansion has been associated with improved access to care and affordability of treatment for medical conditions ranging from colon cancer to end-stage renal disease.^2,3^ However, little is known about the impact of Medicaid expansion on the diagnosis and treatment of melanoma.

Melanoma accounts for the majority of deaths from skin cancer, and its incidence in the United States is increasing.^4^ Melanoma patients who receive early diagnosis and treatment have increased likelihood of survival.^5^ Prior studies have shown that low-income populations have lower rates of early detection and lower rates of survival from melanoma.^6,7^ Therefore, Medicaid expansion could potentially improve these disparities through a number of mechanisms. Medicaid expansion could 1) increase access to primary care providers, 2) increase referrals to dermatologists, 3) reduce delays in seeking care, and 4) reduce the financial burden of melanoma treatment. As such, the aim of this study was to evaluate the effect of Medicaid expansion on changes in insurance status and clinical characteristics of low-income patients with newly diagnosed melanoma.

## Methods

We compared the insurance status of low-income patients with newly diagnosed melanoma residing in Medicaid expansion and nonexpansion states before and after ACA implementation. Our analysis included data from January 1, 2011 to December 31, 2016. 24 states and the District of Columbia expanded their Medicaid programs on January 1, 2014. Therefore, in our analysis, January 1, 2014 marked the beginning of the post-expansion period.

We obtained data from the US National Cancer Institute’s Surveillance Epidemiology and End Results (SEER) program. SEER is a population-based cancer registry that covers approximately 35% of the US population. SEER data collection is standardized to include patient demographics, primary tumor site, stage at diagnosis, and follow-up for vital status.^8^

We considered expansion states to be those that implemented ACA Medicaid expansion in 2014. Within the SEER registry, the expansion states included Alaska, California, Connecticut, Hawaii, Iowa, Kentucky, Michigan, New Jersey, New Mexico, and Washington. The nonexpansion states were Georgia and Utah.

We assembled the study cohort by querying SEER for all malignant melanoma cases (*International Classification of Diseases for Oncology, Third Edition [ICD-O-3]* codes C43.0-43.9) from 2011 to 2016. We first excluded patients greater than or equal to 65 years of age, whom were eligible for Medicare. We excluded cases from Louisiana because Louisiana expanded Medicaid after 2014. We also excluded cases from Massachusetts and New York, where Medicaid was expanded before 2014. We defined our low-income cohort by only including patients who were covered by Medicaid or were uninsured. 97 patients with duplicate records in the registry were excluded from the study cohort.

We described trends in absolute counts and rates of newly diagnosed melanoma patients by insurance status (Medicaid, uninsured) during the pre-expansion and post-expansion periods, stratified by state Medicaid expansion status. We summarized patient level baseline characteristics, including age, sex, race, and reporting source by state Medicaid expansion status. We also described clinical data including total number of tumors per patient, tumor stage, and overall survival. Categorical variables were summarized as counts and percentages and analyzed using the Pearson’s Chi-squared test. Continuous variables were summarized with medians (IQR) and analyzed using the Wilcoxon rank sum test.

We conducted logistic regressions to evaluate the insurance status of newly diagnosed melanoma cases during the pre-expansion and post-expansion periods, stratified by state expansion status. Kaplan Meier curves and Cox proportional hazards model were used to analyze overall survival.

This study did not require IRB approval since the data was de-identified and publicly available. All tests were two-sided with a significance level of 0.05. The analysis was conducted using R software, Version 3.6.2 (The R Foundation for Statistical Computing).

## Results

Our low-income cohort included 4,317 patients with newly diagnosed malignant melanoma, with 3,703 in Medicaid expansion states and 614 in nonexpansion states. Table 1 shows patients from Medicaid expansion states were older than patients from nonexpansion states with a median (IQR) age of 52.00 (42.00, 58.00) vs 49.50 (38.00, 57.00). Other demographic differences were not statistically significant.

**Table 1:**
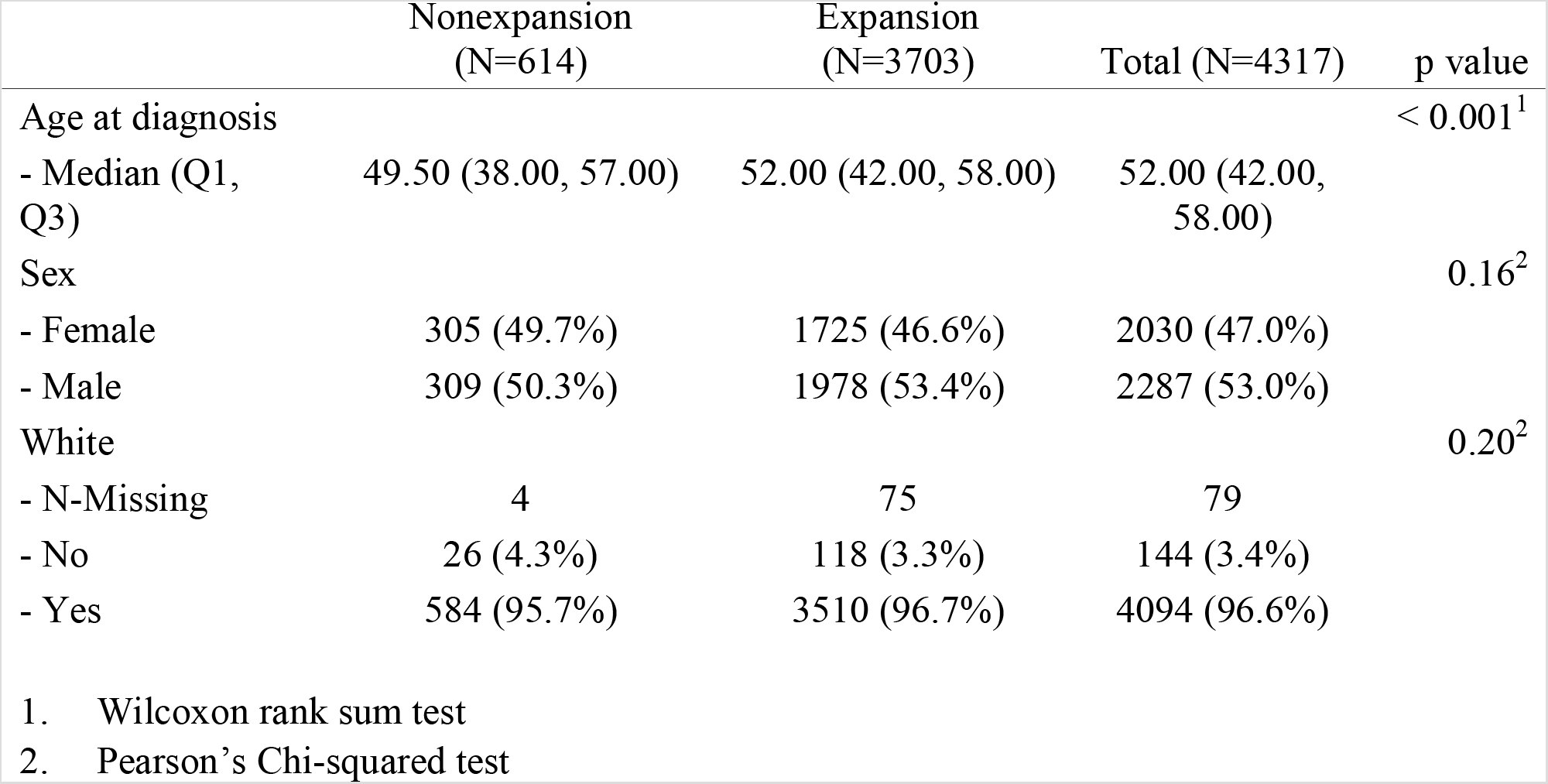
Patient demographics by Medicaid expansion status

In Medicaid expansion states, there were 1,719 low-income patients with newly diagnosed melanoma during the pre-expansion time period (2011-2013) and 1,984 low-income patients with newly diagnosed melanoma (15% increase) during the post-expansion time period (2014-2016). In nonexpansion states, there were 326 low-income patients with newly diagnosed melanoma during the pre-expansion time period, and 288 low-income patients with newly diagnosed melanoma during the post-expansion time period. This difference between Medicaid expansion and nonexpansion states was statistically significant (p< 0.01). (Table 2).

**Table 2:**
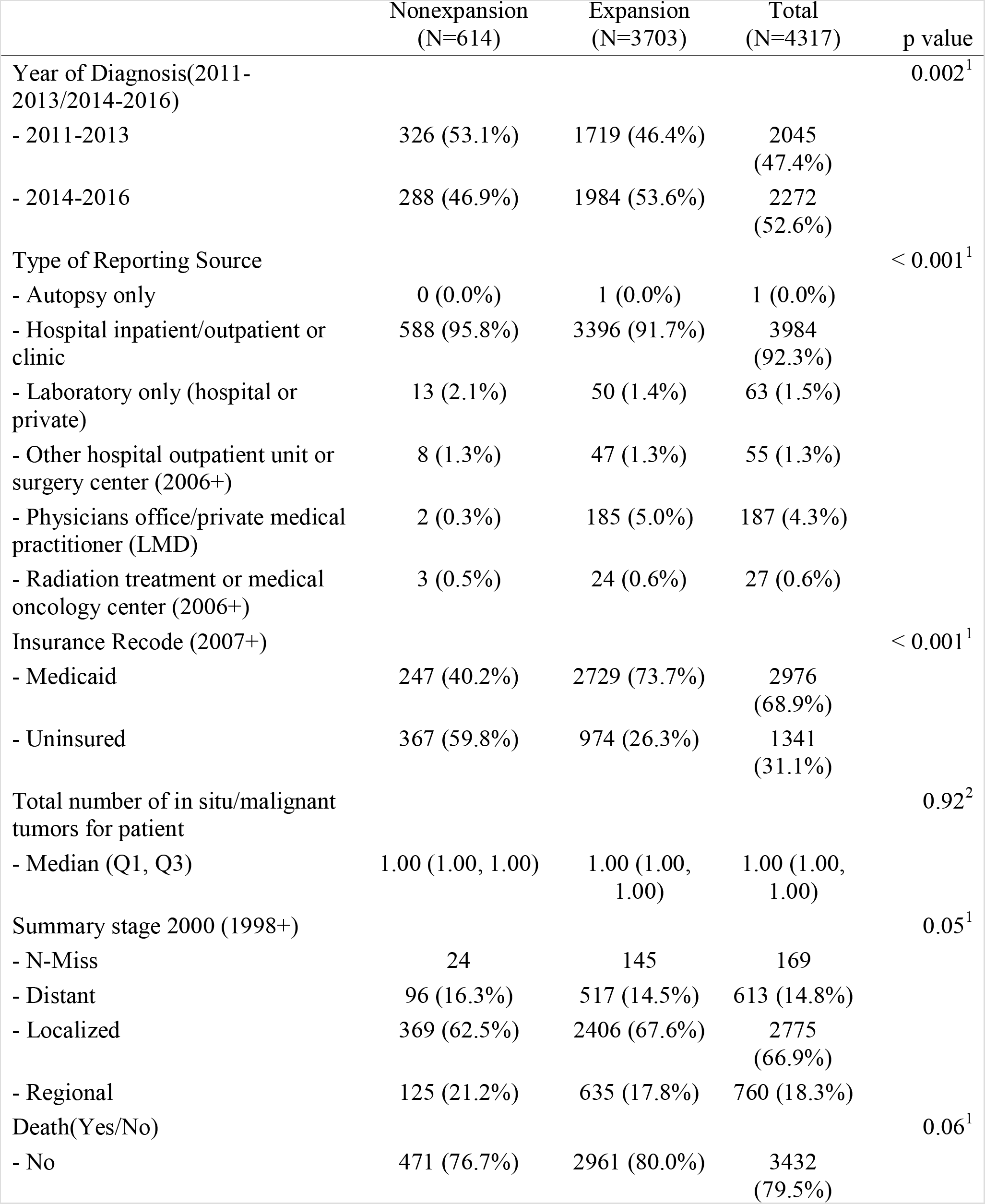
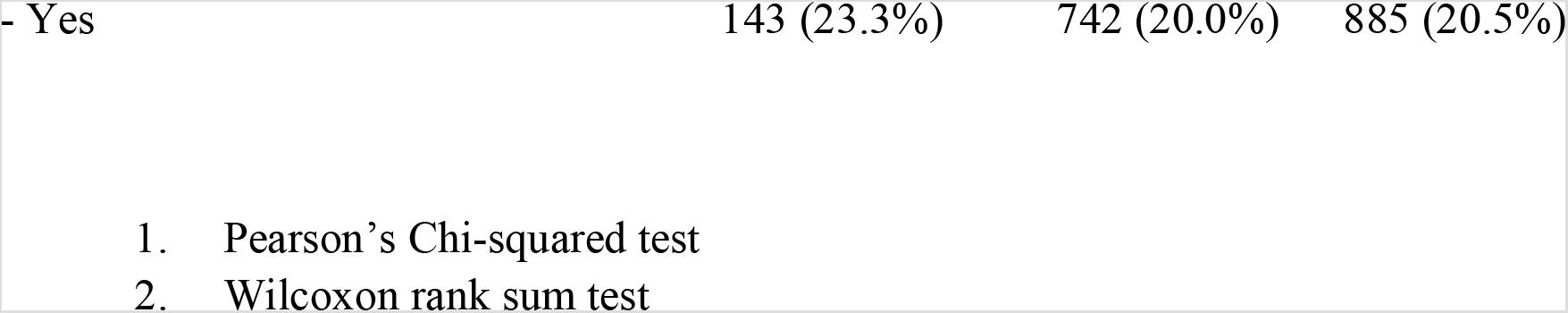
Clinical characteristics by Medicaid expansion status

The clinical characteristics did not differ significantly between patients in expansion versus non-expansion states (Table 2). Overall survival was not statistically different between expansion and nonexpansion states with a HR (95% CI) of 0.89 (0.74, 1.06) (p = 0.20). In Medicaid expansion states, 67.6% of patients had localized melanoma, 17.8% of patients had regional melanomas, and 14.8% of patients had distant melanomas. In nonexpansion states, 62.5% of patients had localized melanomas, 21.2% of patients had regional melanomas, and 16% of patients had distant melanomas. However, these differences between expansion and nonexpansion states were not statistically significant (p = 0.05). Similarly, there were no statistically significant changes in melanoma staging at diagnosis following Medicaid expansion in both the expansion and nonexpansion groups (Table 3).

**Table 3:** Stage at diagnosis by state Medicaid expansion status

| Stage | Non-Expansion<br>(2011-2013) | Non-Expansion<br>(2014-2016) | P value | Expansion<br>(2011-2013) | Expansion<br>(2014-2016) | P value |
| --- | --- | --- | --- | --- | --- | --- |
| Distant | 50(15.9%) | 46(16.7%) | 0.69 | 248(15.1%) | 269(14.1%) | 0.96 |
| Localized | 198(62.9%) | 171(62.2%) |  | 1106(67.2%) | 1300(68.0%) |  |
| Regional | 67(21.3%) | 58(21.1%) |  | 291(17.7%) | 344(18.0%) |  |

**Note:** Chi-Square test

Figure 1 illustrates the insurance status of low-income patients with newly diagnosed melanoma from 2011 to 2016 stratified by Medicaid expansion status. Expansion states had lower rates of uninsured patients throughout the study period. In Medicaid expansion states, the percentage of uninsured patients fell from 36.22% in 2011 to 15.62% in 2016. Over the same time period, the percentage of newly diagnosed patients with Medicaid coverage increased from 63.78% to 84.38%. In nonexpansion states, the percentage of uninsured newly diagnosed patients decreased from 67.96% in 2011 to 58.67% in 2016. Concurrently, the percentage of newly diagnosed patients with Medicaid coverage increased from 32.04% to 41.33%.

**Figure 1:**
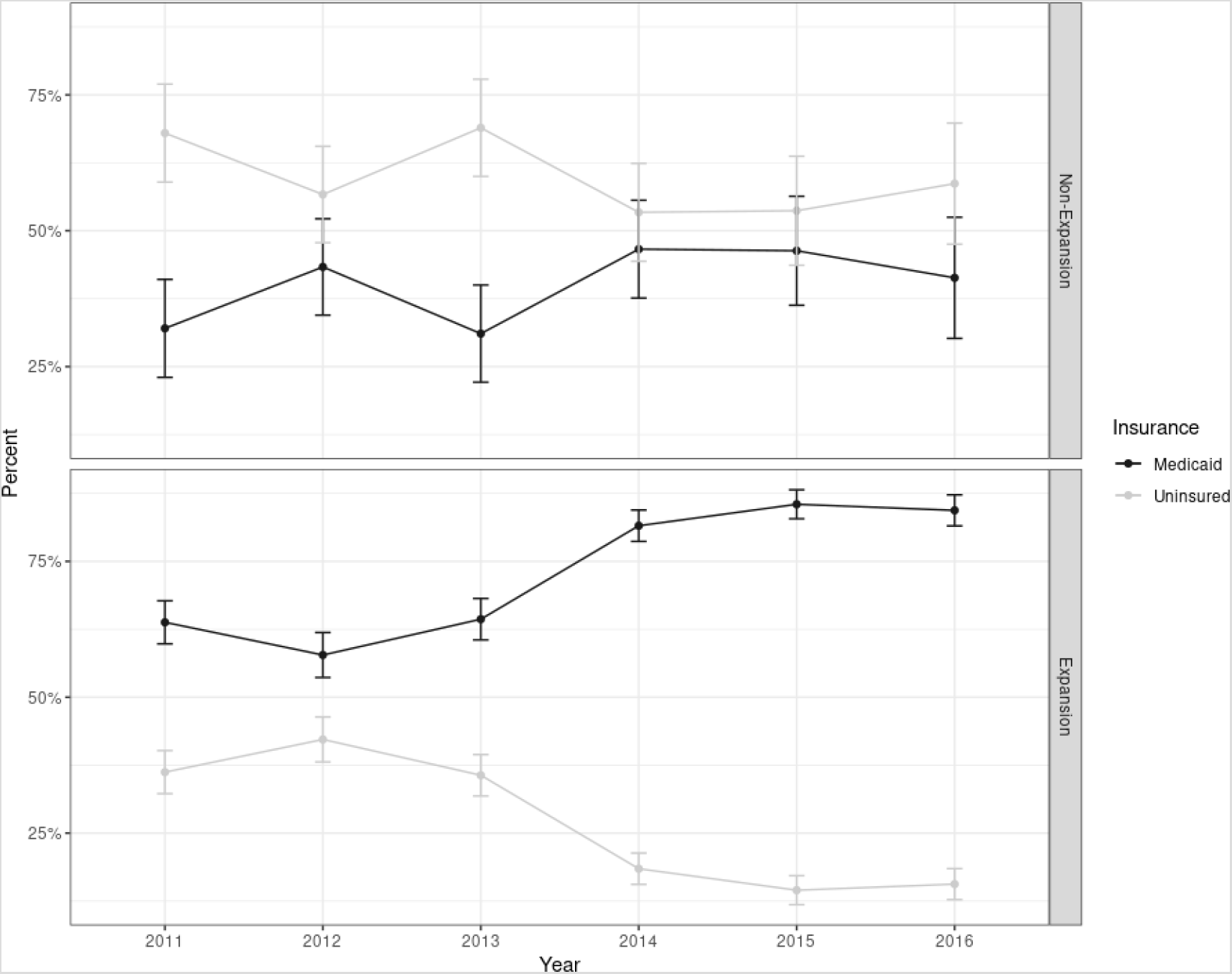
Trends in health insurance status at diagnosis by state Medicaid expansion status

Figure 2 illustrates the adjusted odds ratio of Medicaid insurance status in expansion states compared to non-expansion states for the pre-expansion and post-expansion time periods. During the pre-expansion period, the adjusted odds ratio (95% CI) of expansion states versus non-expansion states was 2.92 (2.28, 3.74) (p< 0.01). In the post-expansion period, the adjusted odds ratio (95% CI) was 6.27 (4.83, 8.14) (p< 0.01). Although the percentage of Medicaid patients increased in both groups following Medicaid expansion, the increase was significantly larger in expansion states as compared to non-expansion states.

**Figure 2:**
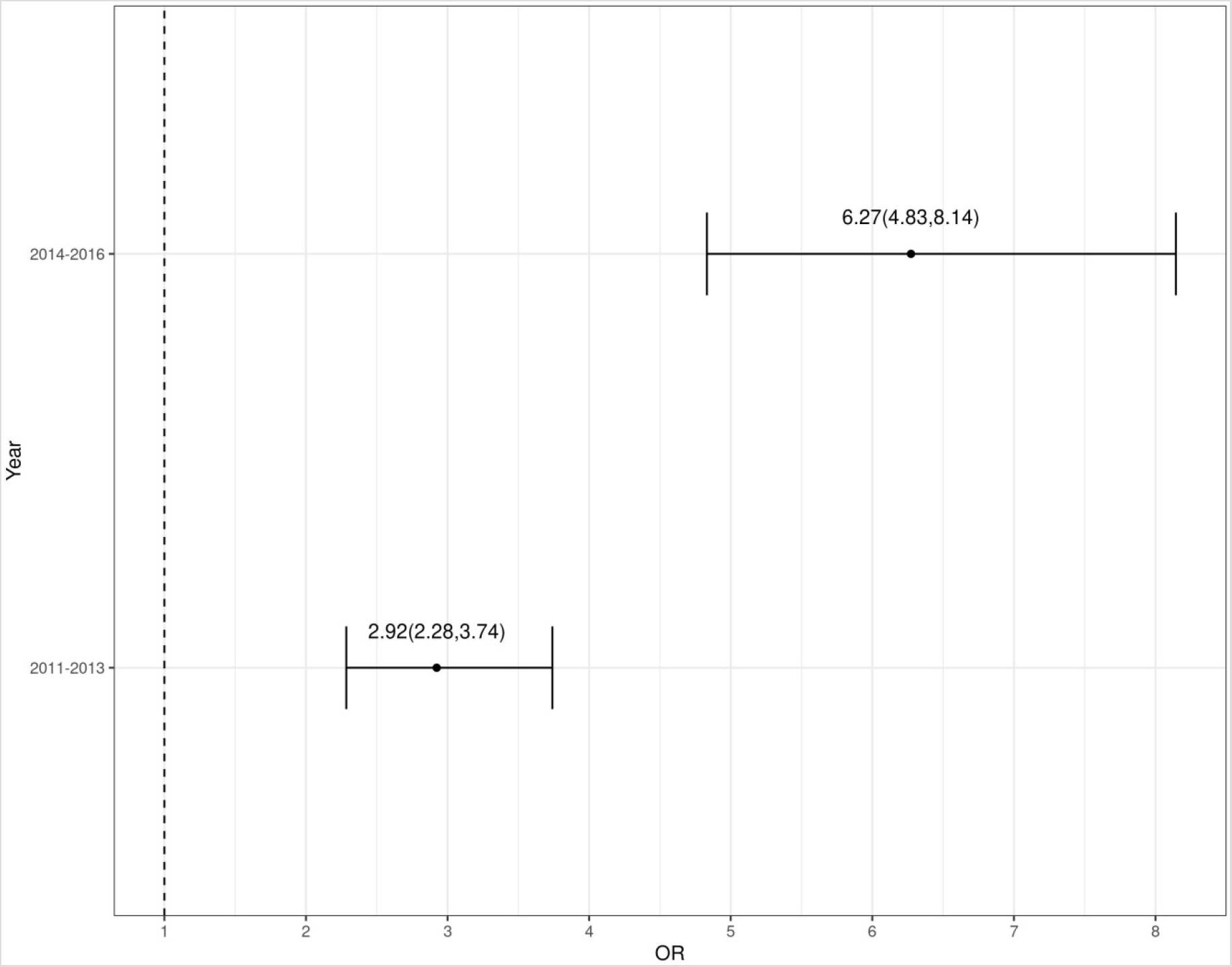
Adjusted odds ratio of Medicaid insurance status in expansion states compared to non-expansion states for the pre-expansion and post-expansion time periods

## Discussion

Using a national population-based cancer registry of newly diagnosed malignant melanoma patients from 2011 to 2016, we found that the ACA’s Medicaid expansion was associated with an increased number of melanoma diagnoses in low-income populations and a decreased rate of uninsured melanoma patients. These findings are consistent with prior studies that evaluated the effects of Medicaid expansion for other cancer patients. Whereas previous studies evaluated the effects of Medicaid expansion only 1 year after ACA implementation, this study encompassed data from 3 years preceding Medicaid expansion to 3 years following Medicaid expansion.^9,10^ Melanoma survival is associated with early treatment, and health insurance coverage has been shown to increase access to high-quality and timely cancer care.^5,11,12^ Therefore, the health insurance coverage gains documented in this study suggest Medicaid expansion could improve timely detection and treatment of melanoma for low-income populations.

In our analysis, Medicaid expansion states had a greater proportion of patients with localized melanoma, while nonexpansion states had a greater proportion of patients with metastatic melanoma. Though these differences were not statistically significant (p = 0.05), a potential explanation for this finding is that increased Medicaid coverage allowed low-income patients to receive earlier diagnoses in Medicaid expansions states. However, in our study, Medicaid expansion states had lower rates of uninsured patients even before 2014, so this finding may reflect underlying differences between expansion and nonexpansion states rather than effects of the ACA implementation.

In addition, prior studies have established wide disparities in melanoma outcomes based on socioeconomic status. Low-income patients present with more advanced melanomas, are at greater risk for surgical delays, and have lower survival rates than higher income patients.^7,13^ Although Medicaid coverage protects low-income patients from the financial toxicities of treatment, it does not guarantee equal access to expert dermatological care. For example, one survey found that only 30% of dermatologists are willing to accept new Medicaid patients.^14^ Furthermore, Medicaid physicians reimbursement rates are, on average, only 72% of Medicare rates.^15^ Therefore, Medicaid’s low physician reimbursement rates in relation to private insurance and Medicare effectively create economic disincentives for physicians to accept new Medicaid patients. Thus, in order to improve timely melanoma treatment for low-income patients, policymakers should not only rely on expanding coverage through Medicaid expansion, but also consider increasing Medicaid reimbursement rates. Though higher physician reimbursement rates may initially increase costs, these costs may be recouped through secondary prevention as patients receive earlier diagnoses and treatment.

This study has several limitations. First, due to its observational design, this study is not able to prove causality. External factors could have changed over time and confounded our temporal comparison of expansion and nonexpansion states. Second, SEER does not provide information on the duration of insurance coverage. Therefore, we could not discern which patients received Medicaid as a direct result of Medicaid expansion versus which patients were already enrolled in Medicaid. Third, insurance status was coded at the time of diagnosis, so our analysis does not capture patients who may have become insured during or after their treatment. Fourth, the SEER database does not include complete national data, so our results may not generalize to states outside of SEER. Fifth, our study was limited to patients younger than 65, and therefore did not include patients who were dual eligible for Medicaid and Medicare.

## Conclusion

The ACA’s Medicaid expansion was associated with increased melanoma diagnoses in low-income patients and a decreased proportion of uninsured patients. However, our study did not identify statistically significant differences in clinical outcomes associated with Medicaid expansion. Therefore, future research is needed to elucidate the long term economic and epidemiological impacts of Medicaid expansion on low-income melanoma patients.

## Data Availability

Data available on request.

## Notes

2 **Conflict of Interest:** The authors report no relevant conflicts of interest

### Competing Interest Statement

The authors have declared no competing interest.

### Funding Statement

No external funding was received to support this work.

### Author Declarations

This study did not require IRB approval because the data used in this study is publicly available and de-identified.

